# A phenome-wide analysis of short- and long-run disease incidence following Recurrent Pregnancy Loss using data from a 39-year period

**DOI:** 10.1101/19003558

**Authors:** David Westergaard, Anna Pors Nielsen, Laust Hvas Mortensen, Henriette Svarre Nielsen, Søren Brunak

## Abstract

**Background:** Pregnancy loss is one of the most frequent pregnancy complications. It is unclear how recurrent pregnancy loss (RPL) impacts disease risk later in life and if later disease risk is different in women with or without a live birth prior to RPL (primary vs. secondary RPL). We sought to investigate if women have an increased risk of disease following RPL, and if there was a difference between primary and secondary RPL.

**Methods:** Using population-wide health care registry databases from Denmark we identified a cohort of 1,370,896 women between 12 and 40 years in the period January 1, 1977, to October 5, 2016 who had been pregnant. Each woman was followed on average for 15.8 years. Of these, 10,691 (0.77%) women fulfilled the criteria for RPL (50.0% had primary RPL). Relative Risk Ratios (RR) were calculated in a phenome-wide manner for diagnoses with a cumulative incidence proportion ≥0.1% in women with RPL. Diagnoses related to assessment and diagnosis of RPL and those appearing later in life were separated using a Mixture Model.

**Results:** In the full cohort of pregnant women, 0.77% (10,691) fulfilled the criteria for RPL (50.0% primary RPL). Compared to women without RPL, primary RPL increased the risk of subsequent cardiovascular disorders, including atherosclerosis (RR=2.45, 1.65-3.51 95% Uncertainty Interval (UI)), cerebral infarction (RR=1.87, 1.43-2.4 95% UI), heart failure (RR=1.97, 1.44-2.63 95% UI), and pulmonary embolism (RR=1.82, 1.32-2.46 95% UI). Women with secondary RPL had an increased risk of obstetric complications, e.g. placenta previa (RR=3.76, 2.9-4.8 95% UI), premature rupture of membrane (RR=2.55, 2.21-2.91 95% UI), intrapartum hemorrhage (RR=2.8, 1.77-4.31 95% UI), gestational hypertension (RR=2.2, 1.67-2.87 95% UI), and puerperal sepsis (RR=2.54, 1.8-3.5 95% UI). We also noticed associations to autoimmune, respiratory, gastro-intestinal and mental disorders in both subtypes.

**Conclusion:** Our findings show that RPL is a risk factor for a spectrum of disorders. This can in part be due to increased screening following RPL, but it also suggests that RPL may directly influence or share etiology with a number of diseases later in life. Research into the pathophysiology of both pregnancy loss and later diseases merits further investigation.

## Introduction

Recurrent Pregnancy Loss (RPL) is a condition that affects 1-3% of couples, depending on its precise definition.^1,2^ While the definition of RPL is not fully consistent across the world, it has historically been defined as three or more consecutive pregnancy losses (PL).^3^ The reason for requiring repeated PLs as part of the RPL definition is that with increasing number of losses the frequency of euploid losses also increases while the chance of a live birth decreases.^4–6^ RPL is divided into two categories: primary and secondary RPL. In primary RPL, there has been no live birth prior to the consecutive losses. In secondary RPL, the women have had at least one live birth. Prior studies have pointed towards a different pathophysiology for these two subtypes. In primary RPL there may be pathophysiological factors that make it impossible to carry a pregnancy to term, whereas in secondary RPL there may be an increased maternal immunization.^11^ Two Scandinavian studies found that 40-48% of cases were secondary RPL.^1,12^ One study from Israel reported that 61% of cases were secondary RPL.^13^

RPL is a multifactorial condition that in addition to fetal causes have multiple known female risk factors such as autoimmune diseases, endocrine disorders, and uterine malformations.^2,9^ Male risk factors have been investigated less, but include chromosomal abnormalities, sperm DNA fragmentation, and age.^10^ Some factors associated with a delayed time to pregnancy and infertility, such as endometriosis, polycystic ovarian syndrome, and diabetes have a clear association to disease later in life, including malignancies, autoimmune diseases, and cardiovascular disease.^14–16^ Moreover, obstetric complications have also been found to increase the risk of cardiovascular disease (CVD).^17^ In recent years, PLs and RPL have been identified as a risk factor for malignancies and cardiovascular disease (CVD).^18–22^ The association with CVD has been further explored in a study that found first degree relatives of women with RPL had an increased risk of CVD.^23,24^ Despite RPL having a major impact on the affected couples, both mentally and physically, the etiology and life-long consequences are poorly understood.^7,8^ Our hypothesis is that RPL and pregnancy losses not attributed to aneuploidy are associated with later disease.

In this study, we compare primary and secondary RPL in a phenome-wide analysis using a nation-wide cohort based on 1,370,896 women.

## Methods

The study was approved by the Danish Data Protection Agency (ref: 2015-54-0939 and SUND-2017-57) and Danish Health Authority (ref: FSEID-00001627 and FSEID-00003092). Informed consent and assessment of the proposal in scientific ethical committees are not required for registry-based research in Denmark.

### Study design and setting

This observational population was identified using the nationwide Danish Patient Registry and the Danish Medical Birth Registry. We included women aged 12-40 with at least one live birth or pregnancy loss (PL) in the period between 1^st^ January 1977 and 5^th^ October 2016. Women with multiple births (twins or higher order) were excluded. The population totaled 1,370,896 women.

Women with RPL were followed from the date of meeting the exposure until the end of follow-up (criteria until death or October 5^th^ 2016).

Each woman with RPL was matched with twenty women from the population without RPL. The women were matched based on year of birth and number of previous live births. The matched comparison group of women without RPL was followed for the outcomes from the same date of the woman with RPL they were matched to, i.e. only the hospital admissions occurring on or after the date of RPL and matching was used in the analysis. Women with RPL were divided into primary and secondary RPL based on the parity history prior to the date of RPL.

Outcomes were identified using several nationwide registries that cover all hospital admissions in Denmark (see Supplementary Methods for a description).^25^

We identified 10,691 women that fulfilled the criteria for RPL (50.0% primary) (defined in the next section). The number of years the women were followed varied based on the outcome, but the total number of person-years was, on average, 1,782,238 years (range 1,231,217-1,799,076 years) for women with primary RPL and the matched group, and 1,744,736 years (range 1,155,427-1,763,733 years) for women with secondary RPL and the matched group. Across all outcomes, the median follow-up up time for women was 15.8 years, ranging from 11.6-16.4 years for each specific outcome.

### Pregnancy Loss and Recurrent Pregnancy Loss

PLs were identified from hospital admissions in the Danish National Patient Registry (Table 1). PLs occurring 8 weeks before or after a molar pregnancy, induced abortion, or extrauterine pregnancy were disregarded (Table 1). In Denmark, every child showing signs of life at delivery is categorized as a live birth irrespective of gestational age. If there is no sign of life at delivery it is considered a pregnancy loss prior to the 28^th^ completed weeks of gestation and a stillbirth hereafter. This definition was changed starting from 2004 when stillbirth were counted from the 22^nd^ completed weeks of gestation. We filtered out diagnoses that were repeated within a medically unreasonable period of time: 1) <22 weeks between two live or still births, 2) <90 days between two PLs, induced abortions, molar or ectopic pregnancies, 3) <22 weeks between a PL, induced abortion, or an extrauterine or molar pregnancy and a stillbirth or livebirth, and 4) <30 days between a live or still birth and a PL, abortion, or an extrauterine or molar pregnancy.

**Table 1:** ICD-8 and ICD-10 codes used to identify recurrent pregnancy loss, pregnancy loss, molar and extrauterine pregnancies. Lowercase x denotes codes including all subcodes.

| Diagnosis | ICD-8/ICD-10 | Surgery | Procedures |
| --- | --- | --- | --- |
| Recurrent<br>Pregnancy Loss | 6430x, Y6439, N96.x, O26.2 | - | - |
| Missed Abortion | 6346x, 6451x, O02.1, O02.1A | - | - |
| Miscarriage | 6438x, 6439x, O03.x | - | - |
| Extrauterine<br>pregnancy | 63109, 6311x, 63129, 63139, 63149,<br>6315x, 63169, 63199, O00.x | 66100, KLBCx,<br>KMCBx, | BKHE0,<br>BKHE8x,<br>BWHA115 |
| Molar pregnancy | 63190, 63429, 63460, 6450x, D39.2,<br>O01.x, O02.0, |  |  |
| Abortion (induced) | 6400x, 6401x, 6402x, 6409x, 6410x,<br>6411x, 6412x, 6413x, 6414x, 6415x,<br>6416x, 6417x, 6419x, 64209, 64219,<br>64229, 64239, 64299, 6455x, 6456x,<br>6458x, O04.x, O05.x, O06.x, O07.x | 63680, 94520,<br>KLCHx,<br>KLWW00, | BKKG1,<br>BKXG1 |

Cases of RPL were identified using hospital discharge codes (Table 1) or by three consecutive PLs. The date of RPL diagnosis was defined as the date of the third PL or RPL diagnosis, whichever came first.

In the matching process, we used 1,370,339 women from the total population. The median number of unique matched women per outcome was 186,752, ranging from 133,875 to 187,355.

### Outcomes

In the phenome-wide analysis we investigated ICD-10 codes at the third level that had a prevalence ≥0.1% in women with RPL (626 diagnoses). ICD-10 codes related to birth, pregnancy loss, and abortion were excluded (O00-O08 and O80-O84), as well as codes from ICD-10 chapters 19, 20, 21, and 22. Only the first instance of a diagnosis code was included.

### Differentiating early from late disease occurrences

Some of the diagnoses with an increased risk are due to investigations at an RPL clinic. To differentiate the diagnoses found during investigations at RPL clinics and later in life, we fitted a log-normal mixture model (LMMM). The optimal number of components was determined using the Bayesian Information Criteria. For each diagnosis code with an increased risk, the time from RPL diagnosis until the outcome occurred was summarized as the median. Time distributions were visualized as histograms that binned multiple diagnoses into one bin.

### Statistical Analysis

Risk ratios (RR) were estimated using a hierarchical Bayesian Poisson log-linear model. As covariates we included the age and previous number of live births. A model was fit separately for women with primary and secondary RPL, respectively. Bayesian posterior distributions were summarized as 95% uncertainty intervals (UI). All estimates were derived from the same Bayesian multilevel model with a pooled prior, which shrinks the expected values and is one way of dealing with the multiple comparison issue.^26^ We defined a prior structure that takes into account the chapter structure in the ICD-10 terminology, see Supplementary Text for a thorough evaluation of the priors and the model. Distributions of time from RPL diagnoses until outcome were modelled using a log-normal mixture model. Additional detail on the statistical procedures are provided in the Supplementary Text. All data were analyzed using stan v2.18, python v2.7 and R v3.1.3.

## Results

Out of a pool of 1,310 unique ICD-10 codes assigned to women after RPL, we investigated 615 diagnoses with a cumulative incidence proportion of at least 0.1% in women with RPL. We found 180 and 172 diagnoses with an elevated RR in primary and secondary RPL, respectively. These covered very heterogenous types of disease (**Figure 1**, Supplementary Table 2), and included cardiovascular disorders, obstetric diagnoses, autoimmune diseases, mental disorders, digestive disorders, and respiratory diseases. We also observed eight diagnoses with a RR lower than one in women with primary RPL, which were all related to complications of labor and delivery.

**Figure 1.**
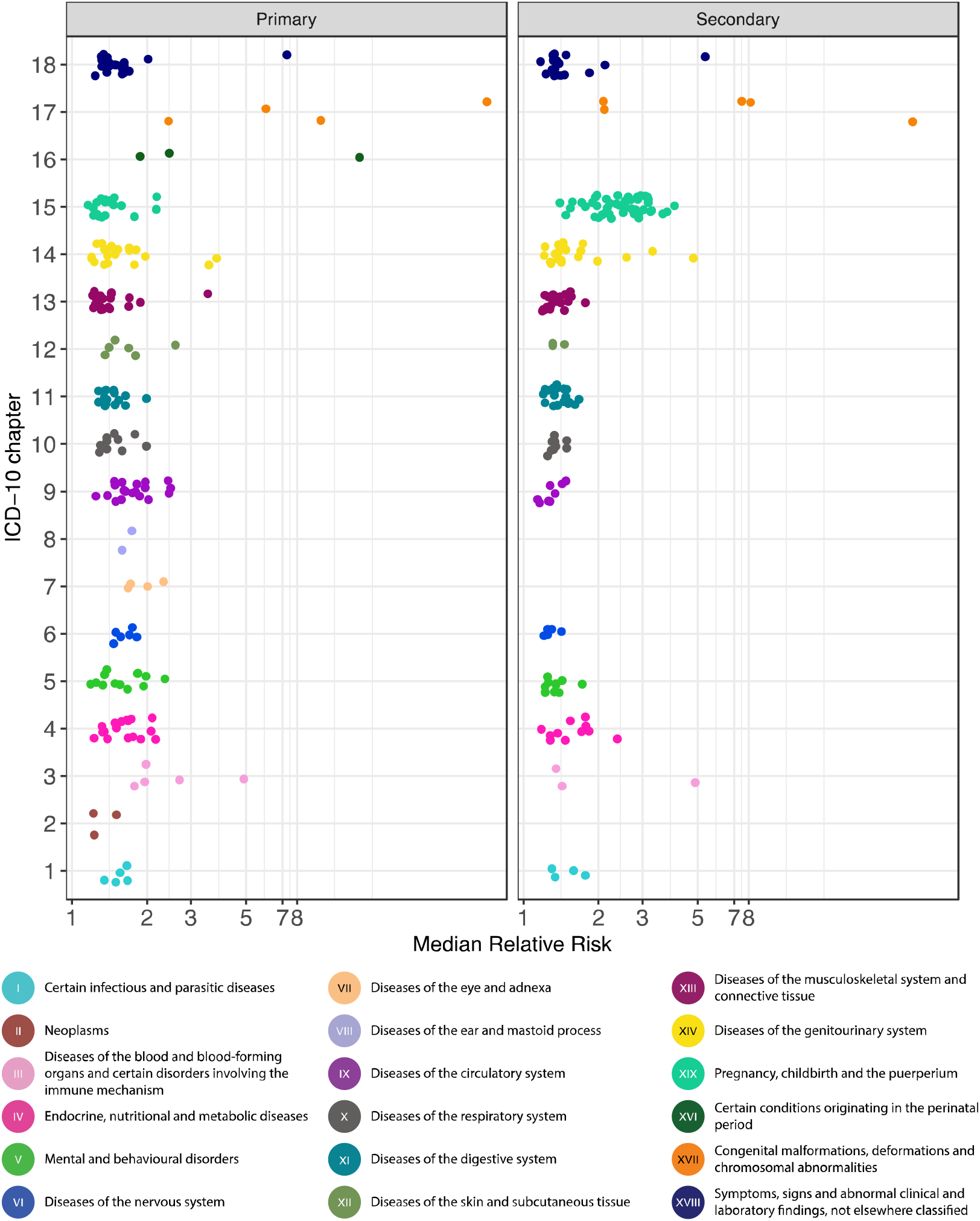
Diagnoses occurring more frequently following RPL across 18 ICD-10 chapters. The points have been scattered in the vertical direction to improve readability. (A) Diagnoses with an elevated risk for women with primary RPL (180 diagnoses). (B) Diagnoses with an elevated risk for women with secondary RPL (172 diagnoses).

Differentiating early from late disease, we identified an “early” and “late” component for both primary and secondary RPL (**Figure 2**, Supplementary Table 3). The early component was found to have a mean of 2.5 (sd=0.9) years for primary RPL and 1.8 (sd=0.7) years for secondary RPL. The component contained a lot of diagnoses routinely investigated as part of the RPL clinic work-up, e.g. uterine and cervical malformations, balanced chromosomal rearrangements, and abnormal immunological findings in serum. Additionally, there were many diagnoses related to obstetric complications and pregnancy for both primary and secondary RPL. When we compared primary and secondary RPL, looking only at the diagnoses with an increased risk in one of the subtypes, we observed many obstetric complications in secondary RPL (Error! Reference source not found.). These included placenta previa (RR=3.76, 95% UI 2.9-4.8), premature rupture of membrane (RR=2.55, 95% UI 2.21-2.91), intrapartum hemorrhage (RR=2.8, 95% UI 1.77-4.31), gestational hypertension (RR=2.2, 95% UI 1.67-2.87), pre-eclampsia (RR=2.31, 95% UI 1.83-2.88), puerperal sepsis (RR=2.54, 95% UI 1.8-3.5), and placental abruption (RR=2.99, 95% UI 2.2-3.97). The latter also had an increased risk in primary RPL, albeit lower (RR=1.32, 95% UI 1.04-1.65).

**Figure 2.**
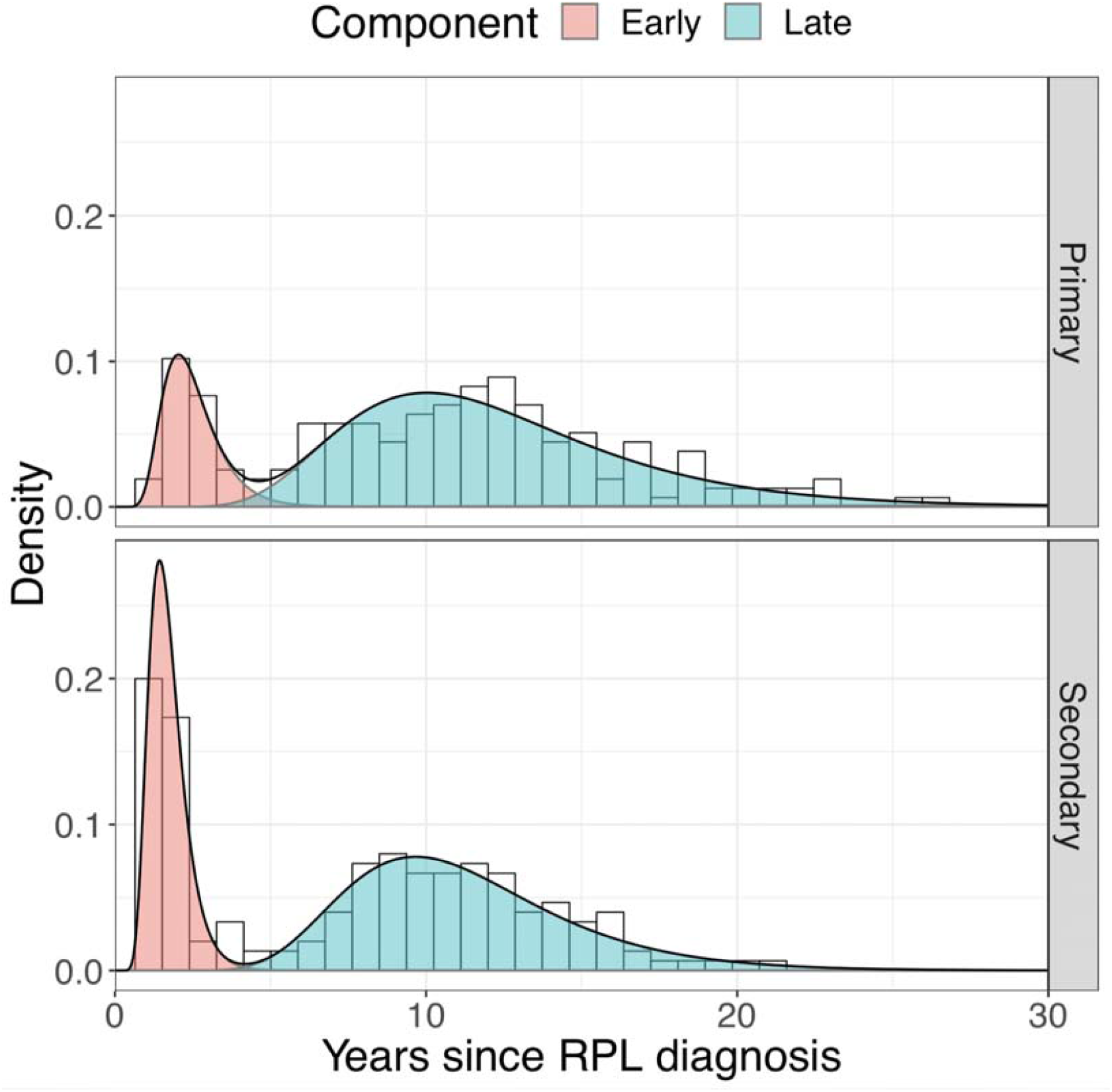
Distribution of the median time between an RPL diagnosis and one of the 180 and 172 diagnoses being more frequent following primary RPL and secondary RPL, respectively. The histogram indicates the observed data points, whereas the two density plots indicate the two components of the mixture model.

**Figure 3.**
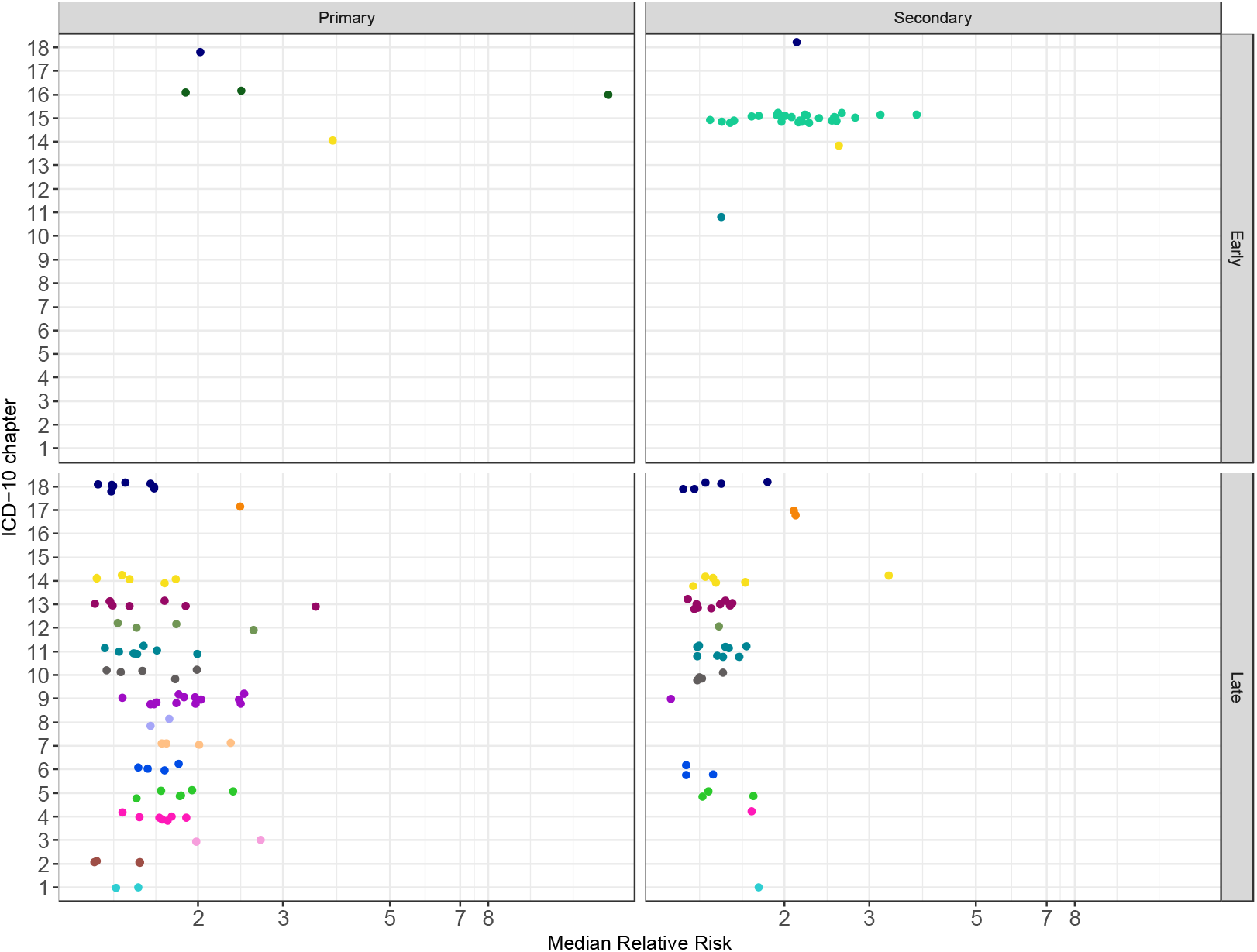
Diagnoses unique to either primary or secondary RPL, divided into the two “early” and “late” groups determined from the mixture model analysis. The coloring scheme is as in Figure 1.

The late component was found to have a mean of 13.3 (sd=5) after the primary PRL diagnosis and contained 160 diagnoses. Strikingly, for many cardiovascular diseases we did not observe an increased risk in women with secondary RPL (Error! Reference source not found.). This included atherosclerosis (RR=2.45, 1.65-3.51 95% Uncertainty Interval (UI)), cerebral infarction (RR=1.87, 1.43-2.4 95% UI), heart failure (RR=1.97, 1.44-2.63 95% UI), and pulmonary embolism (RR=1.82, 1.32-2.46 95% UI). Other diseases included lupus erythematosus (RR=2.6, 1.42-4.53 95% UI), systemic lupus erythematosus (RR=3.51, 2.28-5.21 95% UI), and COPD (RR=1.38, 1.11-1.7 95% UI). Several mental disorders were also found, including anxiety (RR=1.29, 1.08-1.52 95% UI), obsessive-compulsive disorder (RR=1.59, 1.07-2.35 95% UI), and psychiatric disorders associated with the puerperium (RR=2.14,1.15-4.0 95% UI).

In women with secondary RPL, the “late” component had a mean value of 12.0 (sd=3.7) years after the diagnosis. Diagnoses that only had an increased risk in women with secondary RPL included irritable bowel syndrome (RR=1.33, 1.12-1.58 95% UI), ulcerative colitis (RR=1.32, 1.03-1.69 95% UI), and intestinal malabsorption (RR=1.67, 1.16-2.41 95% UI). Mental disorders included recurrent depressive disorders (RR=1.25, 1.07-1.45 95% UI) and mixed personality disorders (RR=1.66, 1.17-2.39 95% UI).

We also observed thyroid disorders (hypothyroidism, nontoxic goiter, hyperthyroidism, and thyroiditis), type 2 diabetes, asthma, reaction to severe stress, depressive episodes, risk of mental disorders due to substance-abuse of alcohol or tobacco, and gastro-intestinal disorders (GERD, gastric ulcer, and gastritis and duodenitis) with an increased risk in both subtypes, belonging to the late component.

## Discussion

In this study, we performed the largest registry-based phenome-wide study to-date of short- and long-run disease incidence associated with recurrent pregnancy loss (RPL). The population comprised 1,370,896 women, of which 10,691 had three or more consecutive PLs. By investigating diagnoses occurring after RPL, we identified distinct spectrums of complications for primary and secondary RPL. The diagnoses were divided into “early” and “late” complications: The “early” complications contained many previously established risk factors for RPL that are part of the routine screening at RPL clinics.^2^ These included coagulation disorders, congenital malformations of the female genital organs, and autoimmune diseases. The component also contained obstetric complications, of which some only had a higher risk in secondary RPL. The “late” component included multiple complications in heterogenous disease domains, such as cardiovascular diseases, autoimmune diseases, mental disorders, digestive system disorders, and respiratory diseases. The disorders occurred, on average, more than ten years after the RPL diagnosis, and there were a distinct set of diseases associated to primary and secondary RPL. We found no evidence that women with RPL had an increased risk of malignancies, irrespective of the subtype.

The basis for this study was nation-wide data collected over a 39-year period. Reporting to the Danish registries used in this study is mandatory. An important source of bias is unregistered PLs. This includes PLs handled at home, in general or gynecological practice. This leads to some women being erroneously classified as not having RPL leading to an underestimation of the RR. Further, the RPL definition depends on the reliability of the identification of spontaneous pregnancy losses in the DNPR. However, this condition has previously been estimated to have a positive predictive value of 97%^27^. Lack of information from the DMBR could result in women being classified wrongly as primary and secondary RPL; nonetheless, the DMBR is considered to be complete.^28^ The registries do not contain complete information on smoking status or BMI. Additionally, we did not have any information on socioeconomic status or whether the partner was the same for all pregnancies, which could confound our findings. Neither did we include the disease history prior to RPL, which could also confound the results. Still, many women experiencing RPL are not properly investigated prior to referral to a specialist clinic, and it is therefore not possible to take this into account. We also note that the infant and maternal mortality in Denmark is extremely low.^30,31^ This is partially due to universal healthcare and referral to specialist clinics involved in the monitoring and treatment of e.g. pregnant women with diabetes and heart disease. Therefore, we would not expect to see many pregnancy losses in the cohort due to e.g. uncontrolled diabetes. Additionally, there are no known causal factors for RPL with the exception of embryonal malformations, and it is thus difficult to control for pre-existing conditions related to RPL. We have attempted to correct for an extremely important factor, namely age, as well as parity which may also affect the health state, as seen with breast cancer.^32,33^ A recent Danish study noted that there is no difference in BMI between women with secondary and primary RPL.^34^ Moreover, the probability of an early pregnancy loss is highly correlated with age. Since we did not have information on the karyotype of the product of conception, we have tried to minimize the confounding effect from aneuploidy by only including women younger than 40 years of age. A mixture of cases due to aneuploidy would weaken the signal, and we could thereby have missed some associations, or underestimated the relative risks. Lastly, this study was based on population data from a 39-year period. Whilst this is helpful for a long follow-up period, changes and developments in medical practice may influence the results. However, we did attempt to mitigate this effect by matching women born in the same year.

Here we studied only the first occurrence of a diagnosis. Nonetheless, it may also be of interest to study if certain conditions have a higher recurrence rate in women with RPL. This is complicated by the fact that it is difficult to determine from the registry whether a disease is actually reoccurring or just a repetition of the existing condition. In our study we used predefined ranges to tackle this issue in relation to birth and pregnancy loss. Lastly, due to the large number of outcomes studied in this phenome wide analysis, we cannot rule out chance findings. Nevertheless, the large number of diagnoses associated with RPL remains striking and is consistent within certain domains of disease, which merits further investigation.

We found that primary and secondary RPL, when considered separate phenotypes, are followed by a unique spectrum of complications. Our observations in the domain of cardiovascular diseases (CVD) and obstetric complications are in accordance with prior studies.^19,20,35,36^ Yet, previous studies of CVD and PL have not been stratified by subtype, and here we demonstrate that primary RPL drives the observed associations. This is an important distinction, as this could point towards shared pathophysiology only present in women with primary RPL and could serve as basis for future screening and risk-assessment profiles. Furthermore, we found that systemic lupus erythematosus (SLE), a recognized risk factor for RPL, also was significantly more frequent in women after a RPL diagnosis and belonged to the “late” component. This can be explained in at least two different ways. First, if SLE and RPL share a common pathophysiological cause our findings indicate that this cause has not always manifested fully at the time of RPL diagnosis and evaluation or it could be that RPL itself increases the risk of SLE possibly through increased microchimerism.^37,38^ Lastly, the increased risk of mental disorders and substance abuse could possibly be mitigated by a closer follow-up and referral to therapy.

The study was performed in a single, national cohort over a period of 39 years. The Danish definition of RPL has been the same in that period, but is different from the one recently adopted by ASRM and ESHRE. Aligning the disease patterns in two cohorts with different definitions can be difficult, and this should be taken into consideration in future comparative studies.

The large population of women, both with and without RPL, allowed us to investigate late-state complication in a phenome-wide manner. This has historically not been possible, owing to the small cohort sizes often used. The findings we present here are of wide interest as we identify RPL as a risk factor and a potential early indicator of later disease, stratified by the subtype.

There are large discrepancies across health in men and women.^39^ Here, we have tried to elucidate how the fertility history contributes to disease. The identification of complications across the full spectrum of disease present a new set of challenges that must be examined in-depth clinically to uncover the etiology. Identification of clinically relevant subtypes is an important aspect of precision medicine, both to tailor screening and therapy to reduce the disease burden, but also to prevent overdiagnosis and unnecessary invasive procedures. Considering the full pregnancy history including prior pregnancy losses is thus relevant when evaluating and predicting the future risk of disease in women.

## Data Availability

Permission to access and analyze data can be obtained following approval from Danish Data Protection Agency and the Danish Health Authority

## Acknowledgements

The work is carried out as a part of the BRIDGE – Translational Excellence Programme (bridge.ku.dk) at the Faculty of Health and Medical Sciences, University of Copenhagen, funded by the Novo Nordisk Foundation (rant agreement NNF18SA0034956). Funding from other Novo Nordisk Foundation grants (NNF14CC0001 and NNF17OC0027594) and the Ole Kirk Foundation and Rigshospitalet’s Research Fund is also acknowledged.

